# DNA sequencing and gene-expression profiling assists in making a tissue of origin diagnosis in cancer of unknown primary

**DOI:** 10.1101/2022.06.24.22276729

**Authors:** Atara Posner, Owen W.J. Prall, Tharani Sivakumaran, Dariush Etemadamoghadam, Niko Thio, Andrew Pattison, Shiva Balachander, Krista Fisher, Samantha Webb, Colin Wood, Anna DeFazio, Nicholas Wilcken, Bo Gao, Christos S Karapetis, Madhu Singh, Ian M Collins, Gary Richardson, Christopher Steer, Mark Warren, Narayan Karanth, Gavin Wright, Scott Williams, Joshy George, Rodney J Hicks, Alex Boussioutas, Anthony J Gill, Benjamin J. Solomon, Huiling Xu, Andrew Fellowes, Stephen B Fox, Penelope Schofield, David Bowtell, Linda Mileshkin, Richard W. Tothill

**Affiliations:** Department of Clinical Pathology and Centre for Cancer Research, University of Melbourne, Melbourne, Australia; Department of Pathology, Peter MacCallum Cancer Centre, Melbourne, Australia; Department of Medical Oncology, Peter MacCallum Cancer Centre, Melbourne, Australia; Sir Peter MacCallum Department of Oncology, University of Melbourne, Melbourne, Australia; Peter MacCallum Cancer Centre, Melbourne, Australia; The Westmead Institute for Medical Research, Sydney, Australia; Department of Gynaecological Oncology, Westmead Hospital, Sydney, Australia; The Daffodil Centre, The University of Sydney, a joint venture with Cancer Council NSW. Sydney, Australia; Department of Medical Oncology, Crown Princess Mary Cancer Centre, Westmead Hospital, Sydney, Australia; Department of Medical Oncology, Flinders University and Flinders Medical Centre, Adelaide, Australia; Department of Medical Oncology, Barwon Health Cancer Services, Geelong, Australia; Department of Medical Oncology, SouthWest HealthCare, Warrnambool and Deakin University, Geelong, Australia; Medical Oncology, Cabrini Health, Melbourne, Australia; Border Medical Oncology, Albury Wodonga Regional Cancer Centre, Albury, Australia; Medical Oncology, Bendigo Health, Bendigo, Australia; Division of Medicine, Alan Walker Cancer Centre, Darwin, Australia; Department of Cardiothoracic Surgery, St Vincent’s Hospital, Melbourne, Australia; Department of Radiation Oncology, Peter MacCallum Cancer Centre, Melbourne, Australia; Department of Computational Sciences, The Jackson Laboratory, Connecticut, United States; The St Vincent’s Hospital Department of Medicine, University of Melbourne, Melbourne, Australia; Department of Medicine, Royal Melbourne Hospital, University of Melbourne, Melbourne, Australia; Cancer Diagnosis and Pathology Group, Kolling Institute of Medical Research and Sydney Medical School, University of Sydney, Sydney, Australia; Department of Psychology, and Iverson Health Innovation Research Institute, Swinburne University, Melbourne, Australia; Behavioural Sciences Unit, Health Services Research and Implementation Sciences, Peter MacCallum Cancer Centre, Melbourne, Australia

**Author notes:** These authors contributed equally to this work (Linda Mileshkin, Richard W. Tothill). Corresponding authors: Richard Tothill, 305 Grattan St, Melbourne VIC 3000, Linda Mileshkin, 305 Grattan St, Melbourne VIC 3000. Competing interest statement: The authors declare no competing financial interests.

**Keywords:** cancer of unknown primary, next-generation sequencing, cancer diagnostic, mutation profiling, targeted therapy, tissue of origin classification

## Abstract

Cancer of unknown primary (CUP) constitutes a group of metastatic cancers in which standardized clinical investigations fail to identify a tissue of origin (TOO). Gene-expression profiling (GEP) has been used to resolve TOO, and DNA sequencing to identify potential targeted treatments; however, these methods have not been widely applied together in CUP patients. To assess the diagnostic utility of DNA and RNA tests for TOO classification, we applied GEP classification and/or gene-panel DNA sequencing in 215 CUP patients. Based on a retrospective review of pathology reports and clinical data, 77% of the cohort were deemed True-CUPs (T-CUP). Among the remaining cases, a latent primary diagnosis (10%) (LP-CUP) or TOO was highly suspected based on combined clinicopathological data (13%) (histology-resolved CUP, HR-CUP). High-medium confidence GEP classifications were made for 80% of LP/HR-CUPs, and these classifications were consistent with a pathologist-assigned diagnosis in 94% of cases, while only 56% of T-CUPs had high-medium confidence predictions. The frequency of somatic mutations in cancer genes was similar to 2,785 CUPs from AACR GENIE Project. DNA features, GEP classification, and oncovirus detection assisted making a TOO diagnosis in 37% of T-CUPs. Gene mutations and mutational signatures of diagnostic utility were found in 31% T-CUPs. GEP classification was useful in 13% of cases and viral detection in 4%. Among resolved T-CUPs, lung and biliary were the most frequently identified cancer types, while kidney cancer represented another minor subset. Multivariate survival analysis showed that unresolved T-CUPs had poorer overall survival than LP/HR-CUPs (Hazard ratio=1.9, 95% CI 1.1 − 3.2, p=0.016), while the risk of death was similar in genomically-resolved T-CUPs and LP/HR-CUPs. In conclusion, combining DNA and RNA profiling with clinicopathological data supported a putative TOO diagnosis in over a third of T-CUPs. DNA sequencing benefited T-CUP tumors with atypical transcriptional patterns that hindered reliable GEP classification.

## Introduction

Cancer of unknown primary (CUP) represents 1-3% of all cancer diagnoses ^1^. It has a notoriously poor outcome, and in Australia, despite being the 14^th^ most common diagnosis is the 6^th^ most common cause of cancer related death ^2^. In the absence of a known tissue of origin (TOO), most CUP patients have historically been treated with empirical chemotherapy that is non-durable in most cases ^3^. Molecularly targeted treatments and immunotherapies may improve the survival outcome for some CUP patients, although drug access can still be challenging without confirming a TOO diagnosis. Furthermore, a CUP diagnosis brings a heavy psychosocial burden, as patients struggle with the absence of a specific diagnosis leading to feelings of high uncertainty ^4^. Therefore, a combination of improved diagnostic and treatment options for people with CUP is urgently needed.

The guidelines for a CUP diagnosis are currently based on standardized gender-appropriate histopathological and clinical investigations, as described by the European Society of Medical Oncology (ESMO) ^5^. Genomic tests, including gene expression profiling (GEP) and DNA methylation analysis, have also been described for making a TOO diagnosis, and some of these tests are commercially available. These classifiers have an accuracy of 83 to 94% when tested on known primary and metastatic tumors ^6–10^ and are superior to immunohistochemistry ^11, 12^. The diagnostic utility of molecular classifiers has been validated when a latent primary can be found (LP-CUP), in which a primary tumor becomes known in time or through alignment with IHC and other clinicopathological information ^6, 9, 11–13^. However, despite the potential diagnostic value of these molecular tests, their clinical utility is questioned. While retrospective and non-randomized studies applying site-directed therapies based on predicted TOO have shown survival benefits ^14, 15^, two randomized clinical trials showed no improvements in patient outcomes ^16, 17^. As such, the level of recommendation for these tests is low under current guidelines ^5^.

DNA mutational profiling has also been explored in CUP ^18–22^. The primary goal of these studies has been to identify actionable mutations to direct targeted therapies with clinically actionable mutations identified in 30-85% of CUP cases ^18–22^. However, mutational profiling also provides insight into the TOO, given that certain mutational processes and the prevalence of cancer driver mutations can be cancer type-dependent ^18^. The availability of large amounts of mutation data in public data repositories enables the interpretation of mutations and mutational signatures found in CUP tumors by a priori knowledge of the frequency and specificity across known cancer types ^23–27^. Not surprisingly, the integration of genomics with histology to resolve CUP diagnosis is making its way into clinical practice ^28^.

While GEP or mutation profiling of CUP has been described independently in previous studies, few studies have considered the diagnostic utility of these assays benchmarked against clinicopathological review. Here we describe the application of GEP and targeted DNA sequencing in a CUP series recruited through a national study (Solving Unknown Primary Cancer: SUPER). Starting from a retrospective review of the available clinicopathological data, we firstly classified CUP cases based on a degree of certainty of a single site TOO diagnosis. We then assessed the potential additive diagnostic value of GEP and DNA sequencing in these patient groups. We also compared overall survival differences and the frequency of potential targeted treatments in the CUP groups based on whether genomics and/or clinicopathological review had assisted in resolving a TOO diagnosis. We used the observations from this study to support the use of genomics data in the diagnostic workup of CUP patients and to further characterize CUP disease characteristics.

## Methods

### Patient cohort

CUP patients were recruited to the SUPER study from 11 Australian sites with informed patient consent under an approved protocol of the Peter MacCallum Cancer Centre (PMCC) human research ethics committee (HREC protocol: 13/62).

Eligibility criteria for patient inclusion in the study were: 1. presenting with carcinoma of no confirmed primary site and who had a preliminary diagnostic workup, including, but not limited to; a detailed clinical assessment; a CT scan of the chest, abdomen, and pelvis; pathological review of tumor tissue; and, other gender-appropriate diagnostic tests; 2. were yet to commence treatment, or had commenced treatment no more than six months prior to recruitment; and 3. could read and write in English, and provide written informed consent. Exclusion criteria included patients under 18 years of age, who had a poor ECOG performance status (ECOG>=3) or uncontrolled medical or psychological conditions that may have prevented completion of study requirements. Patients were identified and referred by their treating clinician at 11 participating hospitals (Supplementary Methods). Histopathology, clinical characteristics, diagnostic investigations, treatment history, and survival data were collected at baseline, six- and twelve-months post recruitment unless deceased or withdrawn earlier.

Latent primary status was recorded by the treating clinician if a primary tumor was detected with both imaging and histopathology during the study. For study purposes, CUP patients were classified by a medical oncologist (TS) into the ESMO defined clinicopathological subsets defined as favorable and unfavorable prognosis groups, as previously described ^5^.

### CUP clinicopathological review

A retrospective review of all histopathology reports and clinical data was performed by a single pathologist (OP) (Supplementary Table 1-2). A histopathology review of slides was undertaken for a subset of cases from the PMCC (n=59), including additional IHC staining if necessary and tissue was available (Supplementary Tables 1-2). Further review of the clinical data was done by a medical oncologist (LM) to concur with the pathologist. Cases were assigned a likely diagnosis where possible, or alternatively, designated as a True-CUP (T-CUP) and then subclassified using a modified version of the Memorial Sloan Kettering Cancer Center OncoTree classification system for CUPs ^29^. OncoTree classification included the following: undifferentiated malignant neoplasms (UDMN), poorly differentiated carcinoma (PDC), adenocarcinoma, not otherwise specified (ADNOS), neuroendocrine tumors, not otherwise specified (NETNOS), neuroendocrine carcinomas, not otherwise specified (NECNOS) and squamous cell carcinomas, not otherwise specified (SCCNOS). ADNOS were further subdivided based on cytokeratin 7 (CK7) and cytokeratin 20 (CK20) IHC staining, and in the case of CK7 negative and CK20 positive staining, caudal-type homeobox 2 (CDX2) was annotated. All ADNOS CK7-CK20+ tumors in the cohort had positive IHC staining of CDX2. SCCNOS were further subclassified to “SCC p16+” based on p16INK4A (p16) IHC staining positivity, as this increases the likelihood of high-risk human papillomavirus (HPV) infection and raises the possibility of oral, uterine, cervical, or anal mucosal origin. The diagnosis for all cases was re-assessed following the NGS findings.

### Gene expression profiling

GEP was performed using a previously described microarray-based test (CUPGuide) ^18^ or a custom NanoString nCounter assay (NanoString Technologies Inc., Seattle, WA, USA). A detailed description of nucleic acid extraction, the NanoString assay and TOO classifier is described in Supplementary Methods. Briefly, the NanoString panel included probe sets targeting 225 genes that were differentially expressed across 18 tumor classes, endogenous control genes, and viral transcripts encoding capsid proteins for HPV16 L1, HPV18 L1, and Merkel cell polyomavirus (VP2) (Supplementary Table 3). The NanoString classifier was trained on harmonized TCGA RNA-seq data previously described ^10^, representing 8,454 samples consolidated into 18 tumor classes (Supplementary Table 4). Tumor classes SCC and Neuroendocrine represented tumors sharing the respective histological features but originating from multiple tissues in training. The RNA-seq/NanoString k-nearest neighbor cross-platform classifier was validated on an independent test set of 188 metastatic tumors profiled by NanoString (see Supplementary Tables 5-6 for further details on the prediction performance and overall accuracy/sensitivity/specificity). A probability score was generated for predictions and heuristic thresholds set for classification confidence level (unclassified <0.5, low ≥ 0.5 and ≤ 0.7, medium confidence >0.7 and < 0.9, high confidence ≥9 probability).

### Targeted DNA sequencing

Targeted enrichment and DNA sequencing were performed on matched blood and tumor DNA, capturing coding regions and exon/intron splice sites of 386 cancer-related genes (listed in Supplementary Table 7) using previously described methods ^30^. Alignment and variant calling were performed using the ensemble variant caller bcbio-nextgen cancer somatic variant calling pipelines and R tools used for analysis. A detailed description of bioinformatics is described in Supplementary Methods.

### Reference mutation data and identification of putative diagnostic DNA features

The AACR Project GENIE mutation data for 77,058 tumors (version 3.7.9) ^23^ was downloaded from the cBioPortal webpage (https://genie.cbioportal.org/) ^26, 27^. The frequency of gene-specific mutations was assessed in tumors annotated as cancer of unknown primary or other cancer types. Genes investigated were restricted to those included in the targeted capture panel in the current study (Supplementary Table 7). For assessment of cancer gene driver mutations, GENIE cancer classes with less than 50 samples per class with genes of interest were removed from consideration, except for assessing gene fusions, where cancer classes with less than 10 samples per cancer class were removed from analysis (Supplementary Table 8).

For identifying DNA features of potential diagnostic utility, the frequency of gene-wise driver mutations was calculated in 22 pre-defined cancer classes (Supplementary Table 8). Oncogenes and tumor suppressor genes (TSGs) were annotated using OncoKB ^24^. For assessing cancer class mutation frequency in the reference data, only truncating mutations in TSGs or hotspot mutations in both oncogenes and TSGs were used (annotated using the web portal http://www.cancerhotspots.org/) ^31^. Oncogenic gene-fusions were restricted to those involving the genes *FGFR2*, *FGFR3*, *ERG*, *FUS*, *TMPRSS2*, *ALK*, *RET*, *ROS1*, *NRG1*, *NTRK1*, *NTRK2*, *NTRK3*, and *EWSR1*. Copy-number alterations were restricted to a curated set of frequently amplified cancer genes (Supplementary Table 7). Gene alteration frequencies was calculated within individual cancer classes (Supplementary Table 9). A Fisher’s exact test was then performed using the R package *stats* to identify genes statistically enriched for DNA alterations in individual cancer classes versus all other cancers. A post hoc adjusted test was performed using the Holm–Bonferroni method. Significance was defined as anything with an adjusted p-value <0.05 and an odds ratio (OR) greater than 1. These significant cancers type diagnostic DNA alterations are summarized in Supplementary Tables 9-10.

### Survival analysis

Overall survival (OS) was measured from the date of CUP diagnosis (histologically confirmed) to the date of death from any cause. Thirty-five patients had death recorded after completion of study follow-up. Patients without a recorded death were censored based on the date of the last follow-up (12 months). Survival analysis was performed using the R package *survival* (v3.1-12). Kaplan-Meier estimates of OS were presented along with log-rank tests for comparison between resolved and true CUP groups. A cox proportional-hazard model was used for multivariable analysis and adjusted for ECOG (0,1,2,3), age (>60 or <60), gender, and ESMO assigned favorable and unfavorable prognosis groups. Kaplan–Meier and forest plots were produced with the *survminer* package (v0.4.8.999).

## Results

### Study cohort and subclassification of CUPs

A total of 215 patients were recruited to the SUPER study, and a summary of baseline characteristics is shown in Table 1. Eighty-nine percent (191/215) received GEP, 93% (201/215) of patients had DNA sequencing, and 82% (177/215) received both assays (Table 1). A latent primary (LP-CUP) cancer was reported by the treating clinician during clinical follow-up in 10% (22/215) of cases based on histopathology, clinical presentation, and/or cancer imaging. In another 13% (27/215) of cases, a likely TOO was assigned after a retrospective review of described morphology, clinical picture, and IHC staining; these cases were termed histology resolved CUP (HR-CUP). Notably, among the LP/HR-CUP cases, there was an enrichment of patients with a prior history of cancer (35%, 17/49), eight of whom likely had a recurrence of their previous disease (Supplementary Table 1).

**Table 1.** Study cohort characteristics

| Characteristics | Count | Proportion |
| --- | --- | --- |
| Total cohort | 215 | 100% |
| Age mean (range) | 61 (20-86) |  |
| Sex |  |  |
| Male | 89 | 41% |
| Female | 107 | 49% |
| Unrecorded | 19 | 9% |
| ECOG |  |  |
| 0 | 67 | 31% |
| 1 | 101 | 47% |
| 2 | 24 | 11% |
| 3 | 1 | 0.5% |
| Not determined | 22 | 10% |
| Previous cancer diagnosis | 58 | 27% |
| ESMO Outcome |  |  |
| Favourable | 31 | 14% |
| Unfavourable | 176 | 82% |
| Not determined | 8 | 4% |
| Gene Expression Profile | 191 | 89% |
| NanoString | 172 | 80% |
| CUPguide | 19 | 9% |
| DNA sequencing | 201 | 93% |
| Both molecular tests* | 177 | 82% |

\*DNA sequencing and GEP performed successfully

Most of the patients (166/215, 77%) were deemed to be True-CUP (T-CUP) and had sufficient IHC workup according to current ESMO guidelines ^5^. Additional IHC stains may have been informative in a small subset (7/166, 4%) but could not be done owing to tissue availability. The T-CUPs were classified into histomorphological subtypes using a modified version of the Memorial Sloan Kettering Cancer Center OncoTree classification (*see* Methods) (Supplementary Table 2). The majority of T-CUPs were adenocarcinomas (51%) or poorly differentiated carcinomas (25%) with minor subsets of SCCs (13%), undifferentiated neoplasms (7%), and neuroendocrine neoplasms (2%). The most frequently assigned cancer types for LP/HR-CUPs or CUP OncoTree classifications are summarized in Supplementary Table 11.

### GEP classification confidence is lower for True-CUPs than known metastatic cancers

We used two GEP methods for TOO classification of 191/215 CUP tumors, where sufficient RNA was available. A previously described 18-class microarray-based classifier (CUPGuide) was used in 20 cases ^18^, while a novel NanoString classifier was used in the remaining cases (Supplementary Table 1-2). The NanoString classifier was validated using an independent cohort of 188 metastatic tumors of known origin (Supplementary Table 6), achieving an overall prediction accuracy of 82.9%, increasing to 91.5% considering only high-medium confidence classifications (n=154) (Figure 1A, Supplementary Table 5.

**Figure 1:**
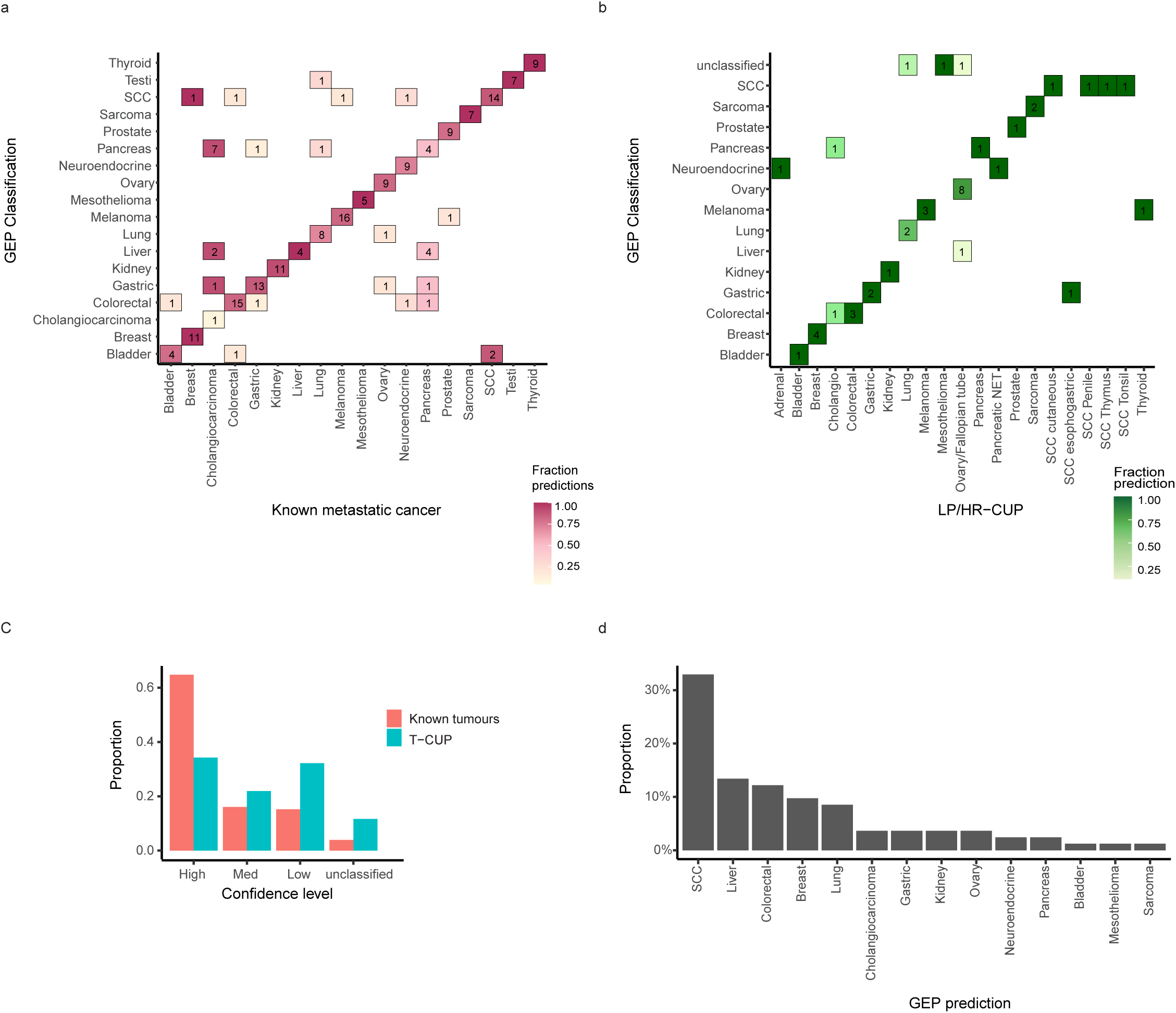
Gene expression profiling (GEP) tissue of origin classification of known metastatic cancers and SUPER cancer of unknown primary (CUP) tumors. A) NanoString GEP classifier was tested on 188 known origin metastatic tumors with confusion matrix showing concordance of tissue of origin prediction and known cancer type. B) GEP classifier tested on latent primary/histology resolved (LP/HR)-CUPs showing concordance between the likely tissue of origin and the predicted cancer type. LP/HR-CUPs representing cancer types that were not represented in the classifier model were removed from analysis C) Fraction of cases within confidence probability score grouping contrasting classification of true-CUPs (T-CUP) and LP/HR-CUPs combined with known metastatic tumors (unclassified <0.5, low ≥ 0.5 and ≤ 0.7 medium confidence ≥ 0.8 and ≤ 0.9, high confidence = 1). D) GEP cancer class predictions of all T-CUPs with high-medium confidence predictions.

GEP TOO classification was possible for 45 LP/HR-CUPs (Figure 1B), of which 80% (36/45) had a high-medium confidence classification (Figure 1C). Three LP/HR-CUPs were considered to be outside the 18-class GEP TOO differential, including one rare ampullary tumor and two uterine tumors. Among high-medium confidence predictions within the GEP diagnostic differential, 94% (31/33) were concordant with their assigned diagnosis (Supplementary Table 1). Common high-medium confidence misclassifications observed among LP/HR-CUPs as well as the 188 known metastatic cancers included cholangiocarcinoma (1/1 and 5/11, respectively) and pancreatic adenocarcinomas (3/10 in the known metastatic group), illustrating classification difficulty among the pancreatobiliary group (Figure 1A, B).

GEP TOO classification was performed on 146 T-CUPs. High-medium confidence classifications were made for only 56% (82/146) of T-CUPs (Figure 1C). The most frequent high-medium confidence classifications included SCC (32%), liver (13%), colorectal (12%), breast (10%) and lung (8.5%) (Figure 1D). A reduced proportion of high-medium confidence classifications for T-CUPs compared to known metastatic cancers suggested T-CUPs either have an atypical transcriptional profile or potentially are cancer types outside of the GEP classifier differential.

### The mutation profile of the SUPER CUP cohort is consistent with other CUP cohorts

DNA panel sequencing was performed for 201/215 CUP tumors and their matching germline controls. We detected mutational features including single nucleotide variants (SNVs), gene-fusions, copy-number amplifications (CNAs), SNV 96 trinucleotide mutational signatures (COSMIC v2)^32^, tumor mutation burden (TMB), and off-target viral DNA sequences (HPV, EBV) (Figure 2). Viral RNA transcripts detected by NanoString also supported viral status for HPV-positive tumors.

**Figure 2:**
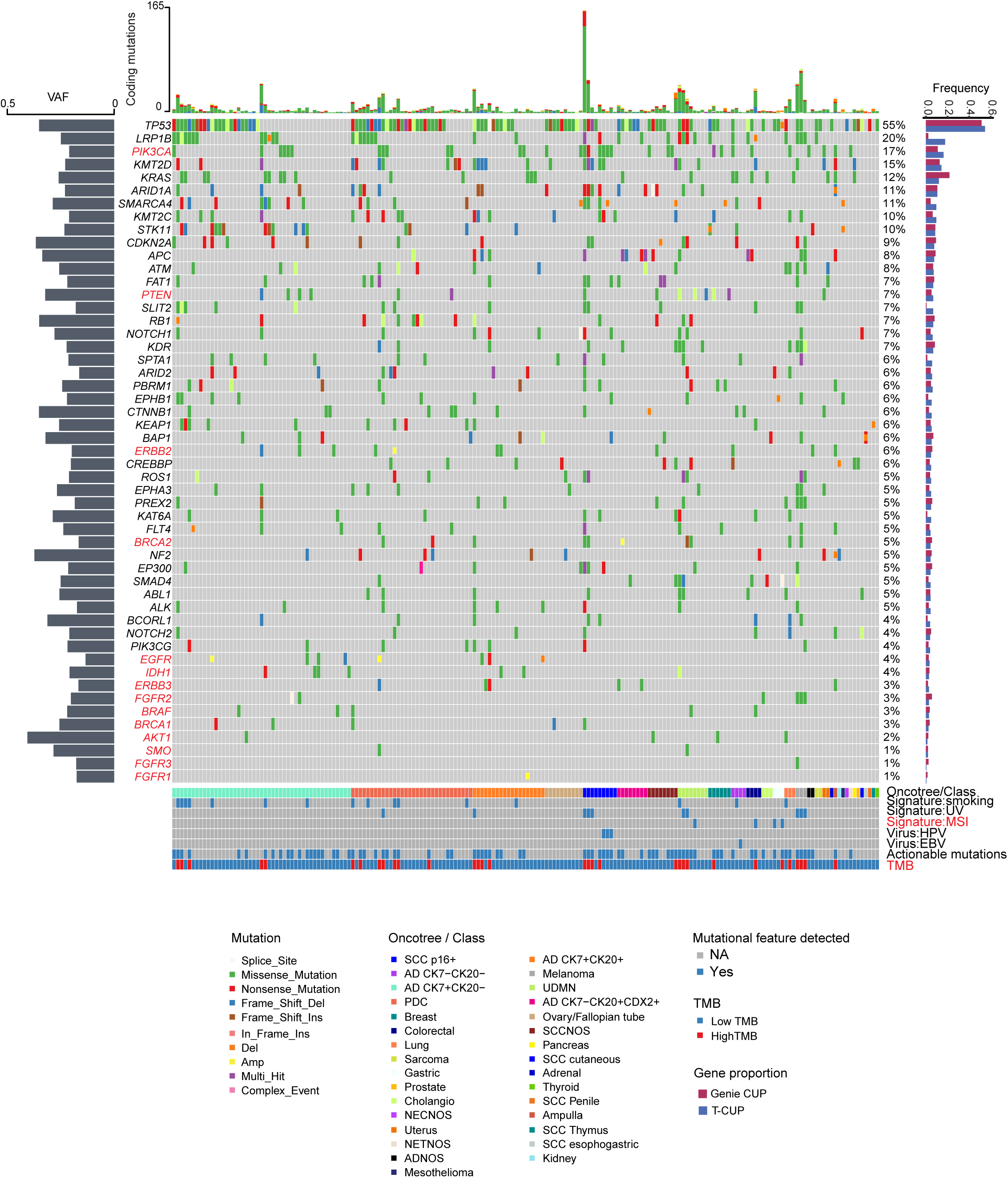
DNA mutational profiling of the SUPER cancer of unknown primary (CUP) cohort. Oncoplot shows somatic mutations in CUPs in descending order of frequency. Genes and mutational features colored in red are actionable and targeted in the CUPISCO trial. The proportion of mutations in the SUPER CUPs was compared with the AACR GENIE CUP cohort (right-hand bar plot). The left-hand plot of the variant allele frequency (VAF) distribution per gene. The top bar plot shows the number of coding mutations per sample. Annotations include OncoTree class for true-CUPs (T-CUP) or the assigned tumor class for latent primary/histology resolved (LP/HR)-CUPs, detection of COSMIC mutational signatures (V2): smoking signature, ultra-violet (UV) signature, DNA mismatch repair signature, oncoviruses: Human papillomavirus 16 (HPV16) and Epstein-Barr virus (EBV), and tumor mutation burden (TMB) status: High >10 mutations/Mb or Low <10 mutations/Mb.

At least one protein-coding mutation was found in 98.5% (198/201) of all CUPs, with a median TMB of 4.4 mutations/Mb (range 0.5-149 mutations/Mb). The most frequently mutated genes were *TP53* (55%), *LRP1B* (20%), *PIK3CA* (17%), *KMT2D* (15%), *KRAS* (12%), ARID*1A* (11%) and *SMARCA4* (11%) (Figure 2). The variant allele frequency of these mutations ranged between 16-40.5%, consistent with clonal cancer driver events when tumor purity was considered. Additionally, 8% (n=16) of the cohort had dominant Signature 4 (tobacco smoking), 5% (n=11) Signature 7 (ultra-violet light, UV) and 2% (n=4) Signature 6 (microsatellite instability, MSI). HPV16 (DNA and RNA) was detected in five cases and EBV (DNA only) in one case.

The gene-wise mutation frequency in the CUP cohort was also compared to 2,785 CUPs with panel sequencing in the AACR Project GENIE database (Figure 2). The mutation profile between the study CUP and GENIE CUP cohorts was highly similar with some minor differences, including a higher frequency of *KRAS* mutations (12% vs. 22%) among the GENIE CUPs and higher *LRP1B* (18% vs. 2%) mutations in the SUPER cohort; the latter explained by *LRP1B* not being included in MSK-IMPACT panel ^33^.

Actionable mutations were also investigated in reference to the CUPISCO trial criteria of actionability described previously ^22^. Eighty-six CUP patients (40%) were matched to either a targeted therapy or immunotherapy arm of CUPISCO (Figure 2) (Supplementary Table 12).

### Mutation profiling and GEP can augment histopathology review

We next considered the diagnostic value of DNA and RNA features, including GEP classification in combination with the histopathological review. Here we considered driver gene mutations, gene fusions, mutational signatures, oncoviruses, and high-medium confidence GEP classifications only. To identify gene features with significant cancer type associations, we referenced the GENIE database (77,058 tumor samples) involving 22 solid cancer types. The potential diagnostic utility of a gene feature was determined by comparing one cancer type versus all others (Fisher exact test adjusted p-value < 0.05 and odds-ratio (OR) >1) (Supplementary Table 10). A total of 171 genes were significantly enriched in one or more cancer types, and 90 genes were enriched in only one cancer type (Figure 3A, Supplementary Figure 1). Mutational signatures also provided cumulative evidence to support likely TOO, for example, a Signature 7 (UV) associated with skin cancer or a Signature 4 (tobacco) found in cancers of the airways, although potentially not excluding liver cancer ^32^.

**Figure 3:**
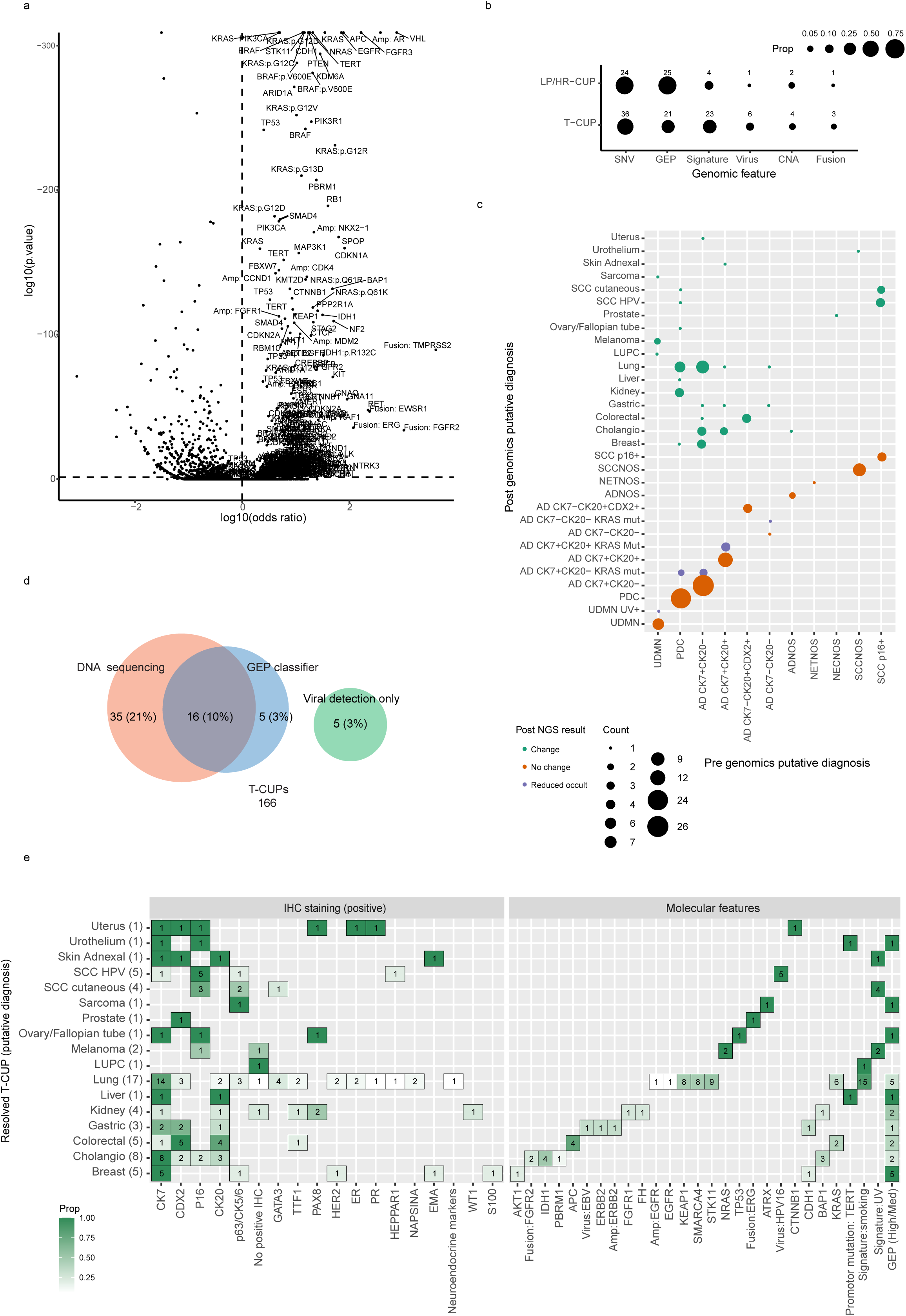
Identification of diagnostically useful DNA features using AACR Project GENIE mutation data and summary of evidence used to support tissue of origin (TOO) diagnosis among SUPER cancer of unknown primary (CUP) cases. A) Volcano plot showing significantly mutated features by cancer types using GENIE pan-caner cohort. All genes are plotted for all cancers using a Fisher’s exact test to calculate adjusted p-values and odds ratio (OR) (significance threshold adjusted p-value < 0.05 and OR >1). B) Proportion and number of cases where each genomic feature supported a putative TOO for true-CUPs (T-CUP) and latent primary/histology resolved (LP/HR)-CUPs. Genomic features included single nucleotide variants (SNV), Gene expression profiling (GEP), mutational signatures, oncoviral sequences, and copy number amplifications (CNA). C) OncoTree cancer classification of T-CUPs before and after genomic analysis. CUPs were either resolved to a putative TOO, had reduced occult diagnosis, or remained T-CUP. D) Proportion of cases where DNA and/or GEP classification supported TOO diagnosis among T-CUPs. E) Detailed summary of supportive genomic features and IHC staining used to assign a putative TOO for all genomically resolved T-CUPs. NGS= Next generation sequencing.

We first assessed LP/HR-CUP cases that had been assigned a single TOO prior to considering the genomics data. Sixty-nine percent of cases (33/49) had one or more features (RNA and/or DNA) consistent with the TOO diagnosis. Both GEP classification and mutation profiling was informative in 20/33 cases, DNA features alone in 7/33, GEP alone in 5/33, and viral detection alone in 1/33 cases. The seven cases where GEP was not informative included two where GEP was not possible, four with low confidence classification, and one was a rare cancer part of the classifier differential. High-medium confidence GEP classifications were the most useful diagnostic feature (n=25) followed by gene mutations (n=24), mutational signatures (n=4), copy number amplifications (n=2), gene-fusions (n=1), and oncoviruses (n=1) (Figure 3B, Supplementary Figure 2).

Examples of LP/HR-CUP diagnoses supported by genomic data included 8/10 (80%) ovarian cases with high-confidence GEP classification as well as a *TP53* mutation, the latter occurring in >96% of high-grade serous ovarian cancers ^34^. A dominant mutation Signature 7 (UV)(n=3) and either oncogenic mutations in *NRAS* (melanoma OR=20.7) or truncating *NF1* mutations (melanoma OR=5.2) was consistent with cutaneous melanoma ^35^. High-confidence GEP prediction of gastric cancer combined with *ERBB2* amplification (concordant with positive HER2 IHC staining) (esophagus/stomach OR=3) and a frameshift deletion in *CDH1* (esophagus/stomach OR=3) supported a diagnosis of gastric adenocarcinoma. High-confidence colorectal GEP prediction and *APC* mutations (colorectal OR= 77.5) were present in two cases diagnosed as colorectal adenocarcinomas ^36^(Supplementary Figure 2, Supplementary Table 1).

A single HR-CUP (1097) had molecular features resulted in a change in classification from colorectal to T-CUP (ADNOS CK7-CK20+CDX2+). No mutations supported a colorectal origin (e.g., *APC*, *RAS/RAF* mutations), and a high-confidence GEP prediction of kidney with mutations in *NF2* and *SMARCA4* was identified. Confirmatory IHC staining (e.g., for PAX8) may have supported a kidney cancer diagnosis, but no tissue was available.

Molecular features supported a likely single TOO diagnosis consistent with clinicopathological features in 37% (61/166) of T-CUPs (Figure 3C). DNA features supported a diagnosis in 31% (51/166) of T-CUP cases, GEP TOO classification in 13% (21/166), and viral detection in 4% (6/166). Combined GEP classification and mutation profiling features were informative in 10% 16/166 cases (Figure 3D). Types of genomic feature supporting a diagnosis included driver gene mutations (n=36), mutational signatures (n=23), HPV16 (n=5) and EBV (n=1) viral nucleic acids, gene amplification (n=4), gene-fusions (n=3) and high-medium confidence GEP prediction (n=21) (Figure 3B). DNA features could narrow the differential diagnosis in a further 11/166 T-CUPs (7%), although a TOO assignment of a single site could not be confidently made (Figure 3C). Considering the genomics data, the most frequently suspected cancer types among T-CUPs were lung (n=18, including a single pleomorphic carcinoma of the lung (LUPC)), biliary tract (n=8), breast (n=5), colorectal (n=5), HPV+ SCC (n=5) and kidney (n=4) (Figure 3E).

Lung-CUP was the single largest single group among T-CUPs. Dominant mutational Signature 4 (tobacco) was found in 14 cases. Driver gene mutations associated with non-small cell lung cancer (NSCLC) included *KEAP1* (Lung OR=9.8), *STK11* (Lung OR=14.1), *SMARCA4* (Lung OR=2.8), and *KRAS* (Lung OR=2.1) (Figure 3E, Supplementary Figure 1 and Supplementary Table 10). Notably, all but one putative Lung-CUP lacked TTF1 IHC staining. Among the Lung-CUPs where GEP was possible, a high-medium confidence classification of Lung was made in 5/16 cases. Two additional T-CUPs that were unresolved had high-medium lung classification, but mutation profiling was unsuccessful, and there was no other evidence to support a lung diagnosis. Three Lung-CUPs were CDX2-positive by IHC but had mutational features consistent with lung cancer, including a Signature 4 (tobacco) in all three cases. GEP was uninformative in these cases as one was classified as colorectal (0.9-confidence probability), and two were predicted SCC, a non-specific classification but potentially in keeping with lung-squamous cancer (Figure 3E, Supplementary Table 2). These CDX2+ Lung-CUPs were favored to be enteric-like lung adenocarcinomas ^37^.

Eight ADNOS tumors were putatively diagnosed as intrahepatic cholangiocarcinomas supported by mutations in *BAP1* (cholangiocarcinoma OR=7.2) and *IDH1* (cholangiocarcinoma OR=32) (Figure 3E and Supplementary Table 10). Seven of these tumors presented with liver masses. These tumors lacked *KRAS* mutations, making a pancreatic origin less likely, given that *KRAS* mutations occur in approximately 90% of pancreatic adenocarcinomas (Supplementary Table 9-10) ^38^. Hotspot *IDH1* (R132C) mutations have the highest frequency in cholangiocarcinoma (cholangiocarcinoma OR=33) but can be detected at a low frequency in melanoma (2%, melanoma OR=3.6). Similarly, *FGFR2-*fusions are frequently detected in intrahepatic cholangiocarcinoma (cholangiocarcinoma OR>100) ^39–41^, a feature that supported a cholangiocarcinoma diagnosis in two T-CUP cases (Supplementary Table 2 and 10).

In four T-CUPs subsequently assigned a kidney TOO, two cases expressed PAX8 by IHC, both confirmed by high *PAX8* mRNA expression (z-score > 2), and an additional case did not have PAX8 IHC performed but had high *PAX8* mRNA expression. The fourth case did not have PAX8 staining or high *PAX8* mRNA expression. Only two cases were classified as kidney cancer by GEP (Figure 3E). Driver mutations significantly associated with kidney cancer, and detected among kidney-CUPs, included *BAP1* (kidney OR=7.6) and *NF2* (kidney OR=7.2). Another case had a truncating *FH* mutation consistent with FH-deficient kidney cancer (kidney OR=8.1) (Supplementary Figures 1). Notably, no *VHL* mutations were detected, representing the most common driver gene in renal cell carcinoma (RCC) (42.5% RCC in GENIE, OR=763). Another assigned kidney case was confirmed as a recurrence of late-onset adult Wilm’s tumor by detecting a somatic *FGFR1* (p.K656D) mutation in both the original primary tumor and a recurrent tumor that presented over twenty years after first diagnosis (Supplementary Table 2 and 10).

### Unresolved CUPs have a reduced survival outcome compared to resolved CUPs

Of the 215 patients, 177 patients had recorded survival data. Among all cases recruited to the SUPER study, the majority were classified as unfavorable according to the ESMO guidelines (137/159= 86% of T-CUP, and 39/48=81% of LP/HR-CUP, where data was available) ^5^ (Figure 4A). Overall, 85% of the cohort was unfavorable CUP and, as expected, these had significantly poorer overall survival (OS)- by log-rank test (p < 0.001), with a median OS of 11 months compared to a median OS of 15 months for favorable CUP. Additionally, 46% of unfavorable CUP achieved 12 months of survival compared with 92% of favorable CUP (Figure 4B). Furthermore, log-rank testing showed longer survival in LP/HR-CUPs, and T-CUPs assigned a single TOO after interpretation of the genomics data, compared with diagnostically unresolved T-CUPs (p=0.04, Figure 4C). Multivariate Cox-regression analysis, including gender, age, ECOG performance, and ESMO prognosis groups (Figure 4D), showed that T-CUPs had worse survival outcomes compared to LP/HR-CUPs (HR=1.89, 95% CI 1.13-3.2, p=0.016) while there was no significant difference between LP/HR-CUPs and resolved T-CUPs (HR=1.18, 95% CI 0.65-2.1, p=0.595).

**Figure 4:**
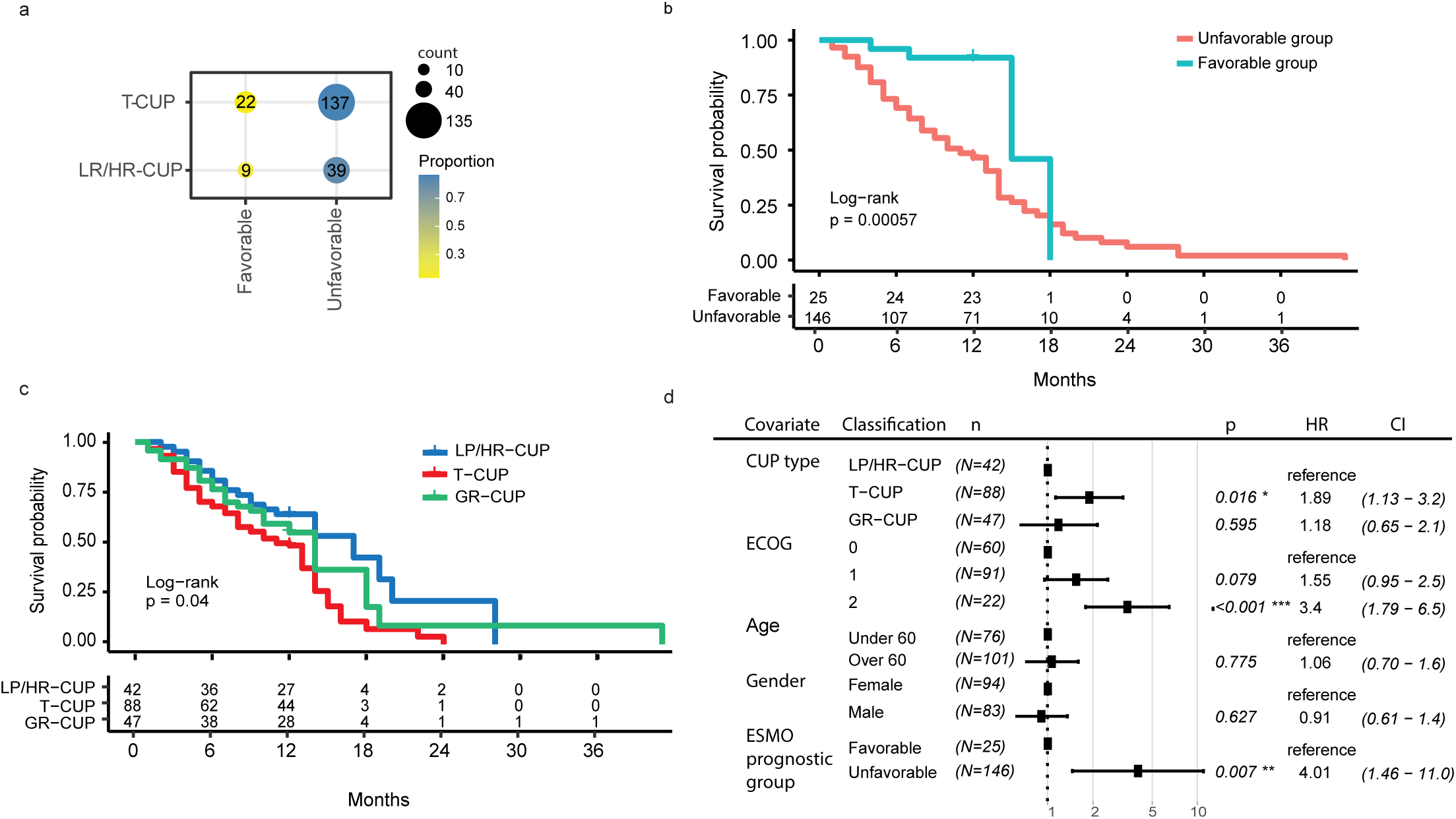
A comparison of overall survival outcome among SUPER CUP patients based on clinical and diagnostic groups A) Proportion and number of true-CUPs (T-CUP) and latent primary/histology resolved (LP/HR)-CUPs classified by European Society of Medical Oncology (ESMO) favorable and unfavorable prognosis groups. B) Kaplan-Meier curve of overall survival (OS) in unfavorable and favorable CUP types. Censored at 12 months and significance using log-rank p-values. C-D) Log-rank test and multivariable Cox-proportional hazard model of SUPER CUP clinical and diagnostic group. Data censored at 12 months and significance using log-rank p values for Kaplan-Meier analysis. Cox-proportional hazard model including covariates of ECOG, age (>60 or <60), ESMO favorable and unfavorable outcome categories, and patient sex. GR = genomically resolved T-CUPs, U= unfavorable type, F= favorable type, HR= hazard ratio, CI= confidence interval.

## Discussion

The reported incidence of CUP has decreased in recent years ^1^. This is most likely due to increased awareness and standardization of investigations, including access to more specific IHC stains, improved diagnostic imaging, and the use of molecular profiling^1^. Consistent with other large retrospective CUP studies, we found that approximately one-quarter of CUP may be assigned a likely TOO based on a centralized histopathology review ^42, 43^. This is similar to the recent experience of the international CUP clinical trial CUPISCO, where ∼20% of patients had a single primary site diagnosis supported by available evidence or was TOO was strongly suspected ^44^. In the current study, we have shown that incorporating DNA and RNA tests may help to resolve more than a third of T-CUP cases not otherwise resolved using conventional testing. Importantly, despite GEP being the most commonly explored molecular test for resolving CUP, we found DNA sequencing was of potentially greater diagnostic value among CUP tumors due to many CUPS having an atypical transcriptional profile.

Despite positive evidence that GEP can be diagnostically informative for CUP, cases with poorly differentiated histopathology and atypical disease presentation remain challenging ^45, 46^. This is perhaps not surprising given that GEP classification relies upon the expression of cellular differentiation markers that are often lost or equivocal in CUP tumors ^47^. Our observation that fewer CUP pass a high-medium confidence GEP classification compared to known metastatic cancers, hence have a reduced classifier performance, is supported by other studies. For instance, the CancerTYPE ID (Biotheranostics) 92-gene test, which had extensive multisite validation, showed an overall accuracy of 85% for known cancer metastases. But in CUP, the concordance of GEP with IHC and clinicopathological evidence was lower at 75% for LP-CUP and 70% compared to the clinical picture only ^7^. Additionally, rare malignancies are infrequently included in the training set of TOO classifiers ^6^, which poses a limitation for rare entities among a cohort of CUP. More specifically, subsets of lung and biliary cancers can be problematic for GEP classifiers to resolve. For example, among lung CUPs, GEP classification was found to be concordant with a latent primary tumor in only 50% of cases ^13^. GEP classification accuracy can also be low for cholangiocarcinomas, as the transcriptional profile is similar to pancreatic and upper gastrointestinal neoplasms ^10^. Notably, some previously described GEP and DNA methylation tests have also excluded cholangiocarcinoma in their models or combined them into a pancreaticobiliary class ^9, 48^. We found that DNA sequencing may be particularly useful in pancreaticobiliary CUPs given that gene mutations characteristic of cholangiocarcinomas can have both diagnostic and occasionally therapeutic significance, including alterations in *IDH1*, *FGFR2,* and *BAP1* ^39–41^.

Integrated molecular profiling of CUP can help identify rare disease subtypes and our data supports their being some recurrent CUP entities. We identified examples of pulmonary enteric adenocarcinomas that lack TTF1 expression but can express the gastrointestinal marker CDX2 ^37^. GEP or IHC alone could not resolve such cases, given their atypical profile; however, they still retained DNA features highly suggestive of NSCLC, including a tobacco mutational signature and somatic mutations in *KRAS*, *STK11*, and *SMARCA4*. *SMARCA4*-deficient lung cancers are known to lack TTF1 expression ^49^ and are likely to be frequent in the CUP population ^19, 20^. *SMARCA4*- deficient lung cancer models similarly lose expression of lung differentiation markers and have a pro-metastatic behavior consistent with aggressive clinical course and poor survival outcomes in patients ^50^. Kidney cancers are another emerging CUP entity representing ∼4-6% of CUPs ^44, 51, 52^. Four CUPs in the SUPER cohort were consistent with a primary kidney tumor. Interestingly, we found somatic mutations in *NF2*, *FH*, and *BAP1* in some RCC cases. Somatic *NF2* mutations are characteristic of advanced papillary renal cell tumors and those with biphasic hyalinizing psammomatous features ^53^. Papillary carcinomas have also been found to be enriched among other Kidney-CUPs ^51, 52^ with a mutation profile similar to the Kidney-CUPs described in the SUPER cohort ^54^. Identifying CUP entities and recurrent therapeutic targets in these groups may help guide future CUP clinical trials. For instance, while empirical chemotherapy is ineffective in RCC, targeted therapies and ICIs are likely more efficacious ^51, 55^. Furthermore, detection of *NF2* mutations among RCC and mesothelioma could direct targeted treatment of the Hippo pathway using inhibitors of TEAD auto-palmitoylation ^56^, which have now entered clinical trials (NCT04665206).

CUP can be classified into favorable and unfavorable prognosis groups. Given that ∼85% of CUPs belong to the unfavorable group, research and clinical trials directed at identifying other treatments for these patients or therapeutic responsive subsets should be sought. Approximately 40% of T-CUPs in the SUPER cohort had potentially actionable mutations and genomic alterations, broadly consistent with other CUP profiling studies ^19, 20, 22^. As curated findings from genomic tests were returned to clinicians during the study, it is possible that the genomic data allowed site-specific treatments and access to targeted treatments, potentially explaining the improved survival outcome among the resolved compared to the unresolved T-CUP group. Alternatively, genomically resolved CUPs may be enriched for cancer types where targeted or immunotherapy treatments are available and effective.

In conclusion, we have shown that DNA and RNA tests can be incorporated into a pathology assessment to improve cancer type diagnosis, identify CUP subtypes, and direct treatments. Rather than replacing traditional histopathological analysis, molecular testing can augment conventional testing to either confirm a suspicion of primary tissue of origin or provide robust diagnostic leads that are not otherwise evident using other modalities. In practice, in cases where tissue is limited, it may be more informative to prioritize genomic testing to guide additional investigations before consuming tissue on extended IHC panels. With steady improvements in technology and reduction in the sequencing costs, more comprehensive whole-genome and transcriptome analysis will likely increase the sensitivity to detect features such as structural variants and mutational signatures that are not reliably detected by panel sequencing ^57^. Recent studies have also utilized machine-learning for classification based on genome-wide patterns of somatic mutations that are not otherwise interpretable by human curation ^58, 59^ and combined DNA and RNA features ^60^ to improve the TOO prediction accuracy further (60-91% and 94%, respectively). Although molecular testing is not currently recommended in most CUP guidelines, careful prospective evaluation of these tests may justify their cost, helping to resolve cancer type in a significant fraction of CUP cases as well as direct treatments that will ultimately translate to better outcomes for patients.

## Data Availability

The datasets used and/or analyzed during the current study are available from the corresponding author on reasonable request.

## Supplementary Information

### Supplementary Methods

**Supplementary Figure 1:** Diagnostically useful mutated features per cancer type using GENIE pan-caner cohort.

**Supplementary Figure 2**: Supportive genomic features and IHC staining for latent primary and histology resolved CUPs.

Supplementary Tables 1-12

## Acknowledgments

We acknowledge Cameron Patrick of the Melbourne Statistical Consulting Platform for providing statistical support. We wish to thank Jillian Hung and Niklyn Nevins, SUPER study coordinators at Westmead and Blacktown, Lisa Kay at Nepean, Karin Lyon (ethics and governance), and acknowledge the contributions of the Nepean Cancer Biobank and the Westmead GynBiobank for facilitating the study. The authors would like to acknowledge the American Association for Cancer Research and its financial and material support in the development of the AACR Project GENIE registry, as well as members of the consortium for their commitment to data sharing. Interpretations are the responsibility of study authors. We wish to acknowledge the patients who have contributed to this study and the CUP consumer steering committee, Cindy Bryant (chair), Kym Sheehan, Christine Bradford, Clare Brophy, Dale Witton, and Frank Stoss.

## Ethics Approval / Consent to Participate

All participating patients provided informed consent to partake in the study. This was approved by the Peter MacCallum Cancer Centre (PMCC) human research ethics committee (HREC protocol: 13/62).

## Author Contribution Statement

RT and LM conceived the study. AP performed the analysis, drafted the figures and tables. OWJP undertook the histopathology review and data analysis. TS and LM reviewed the clinical data. CW, KF, SW collected the data for the study. ADF, NW, CSK, BG, CS, MS, IMC, GR, MW, NK screened patients for eligibility. DE and NT developed the NanoString classifier, and JG and SB validated the classifier. GW, SW, AB, AJG, BJS, DB, SF, RJH contributed samples for training the classifier. AF, HX and SF performed the mutation profiling. APa performed the bioinformatic analysis. AP and RT co-wrote the manuscript. All authors critically reviewed the manuscript. PS, DB, LM and RT are the principal investigators and obtained research funding to support the study.

## Reference List

1 Rassy, E. & Pavlidis, N. The currently declining incidence of cancer of unknown primary. Cancer Epidemiol 61, 139–141 (2019).

2 Australian Institute of Health and Welfare (AIHW) 2018Cancer Data in Australia; Australian Cancer Incidence and Mortality (ACIM) books: cancer of unknown primary site Canberra: AIHW.

3 Massard, C., Loriot, Y. & Fizazi, K. Carcinomas of an unknown primary origin--diagnosis and treatment. Nat Rev Clin Oncol 8, 701–710 (2011).

4 Hyphantis, T., Papadimitriou, I., Petrakis, D., Fountzilas, G., Repana, D., Assimakopoulos, K. et al. Psychiatric manifestations, personality traits and health-related quality of life in cancer of unknown primary site. Psycho-oncology 22, 2009–2015 (2013).

5 Fizazi, K., Greco, F. A., Pavlidis, N., Daugaard, G., Oien, K., Pentheroudakis, G. et al. Cancers of unknown primary site: ESMO Clinical Practice Guidelines for diagnosis, treatment and follow-up. Ann Oncol 26, v133–138 (2015).

6 Erlander, M. G., Ma, X. J., Kesty, N. C., Bao, L., Salunga, R. & Schnabel, C. A. Performance and clinical evaluation of the 92-gene real-time PCR assay for tumor classification. J Mol Diagn 13, 493–503 (2011).

7 Kerr, S. E., Schnabel, C. A., Sullivan, P. S., Zhang, Y., Singh, V., Carey, B. et al. Multisite validation study to determine performance characteristics of a 92-gene molecular cancer classifier. Clin Cancer Res 18, 3952–3960 (2012).

8 Hainsworth, J. D. & Greco, F. A. Gene expression profiling in patients with carcinoma of unknown primary site: from translational research to standard of care. Virchows Arch 464, 393–402 (2014).

9 Moran, S., Martínez-Cardús, A., Sayols, S., Musulén, E., Balañá, C., Estival-Gonzalez, A. et al. Epigenetic profiling to classify cancer of unknown primary: a multicentre, retrospective analysis. Lancet Oncol 17, 1386–1395 (2016).

10 Zhao Y, P. Z., Namburi S, Pattison A, Posner A, Balachander S, Paisie C.A., Reddi H.V., Rueter J, Gill AJ, Fox S, Raghav KPS., Flynn WF, Tothill RW, Li S, Karuturi RKM, George J. CUP-AI-Dx: a tool for inferring cancer tissue of origin and molecular subtype using RNA gene-expression data and artificial intelligence. EBioMedicine (2020).

11 Handorf, C. R., Kulkarni, A., Grenert, J. P., Weiss, L. M., Rogers, W. M., Kim, O. S. et al. A multicenter study directly comparing the diagnostic accuracy of gene expression profiling and immunohistochemistry for primary site identification in metastatic tumors. Am J Surg Pathol 37, 1067–1075 (2013).

12 Tothill, R. W., Shi, F., Paiman, L., Bedo, J., Kowalczyk, A., Mileshkin, L. et al. Development and validation of a gene expression tumour classifier for cancer of unknown primary. Pathology 47, 7–12 (2015).

13 Greco, F. A., Lennington, W. J., Spigel, D. R. & Hainsworth, J. D. Molecular profiling diagnosis in unknown primary cancer: accuracy and ability to complement standard pathology. J Natl Cancer Inst 105, 782–790 (2013).

14 Hainsworth, J. D., Rubin, M. S., Spigel, D. R., Boccia, R. V., Raby, S., Quinn, R. et al. Molecular gene expression profiling to predict the tissue of origin and direct site-specific therapy in patients with carcinoma of unknown primary site: a prospective trial of the Sarah Cannon research institute. J Clin Oncol 31, 217–223 (2013).

15 Yoon, H. H., Foster, N. R., Meyers, J. P., Steen, P. D., Visscher, D. W., Pillai, R. et al. Gene expression profiling identifies responsive patients with cancer of unknown primary treated with carboplatin, paclitaxel, and everolimus: NCCTG N0871 (alliance). Ann Oncol 27, 339–344 (2016).

16 Haratani, K., Hayashi, H., Takahama, T., Nakamura, Y., Tomida, S., Yoshida, T. et al. Clinical and immune profiling for cancer of unknown primary site. J Immunother Cancer 7, 251 (2019).

17 Fizazi, K., Maillard, A., Penel, N., Baciarello, G., Allouache, D., Daugaard, G. et al. LBA15_PRA phase III trial of empiric chemotherapy with cisplatin and gemcitabine or systemic treatment tailored by molecular gene expression analysis in patients with carcinomas of an unknown primary (CUP) site (GEFCAPI 04). Annals of Oncology 30 (2019).

18 Tothill, R. W., Li, J., Mileshkin, L., Doig, K., Siganakis, T., Cowin, P. et al. Massively-parallel sequencing assists the diagnosis and guided treatment of cancers of unknown primary. J Pathol 231, 413–423 (2013).

19 Ross, J. S., Wang, K., Gay, L., Otto, G. A., White, E., Iwanik, K. et al. Comprehensive Genomic Profiling of Carcinoma of Unknown Primary Site: New Routes to Targeted Therapies. JAMA Oncol 1, 40–49 (2015).

20 Varghese, A. M., Arora, A., Capanu, M., Camacho, N., Won, H. H., Zehir, A. et al. Clinical and molecular characterization of patients with cancer of unknown primary in the modern era. Ann Oncol 28, 3015–3021 (2017).

21 Löffler, H., Pfarr, N., Kriegsmann, M., Endris, V., Hielscher, T., Lohneis, P. et al. Molecular driver alterations and their clinical relevance in cancer of unknown primary site. Oncotarget 7, 44322–44329 (2016).

22 Ross, J. S., Sokol, E. S., Moch, H., Mileshkin, L., Baciarello, G., Losa, F. et al. Comprehensive Genomic Profiling of Carcinoma of Unknown Primary Origin: Retrospective Molecular Classification Considering the CUPISCO Study Design. Oncologist (2020).

23 Consortium, A. P. G. AACR Project GENIE: Powering Precision Medicine through an International Consortium. Cancer Discov 7, 818–831 (2017).

24 Chakravarty, D., Gao, J., Phillips, S., Kundra, R., Zhang, H., Wang, J. et al. OncoKB: A Precision Oncology Knowledge Base. JCO Prec Oncol 1–16 (2017).

25 Forbes, S. A., Beare, D., Boutselakis, H., Bamford, S., Bindal, N., Tate, J. et al. COSMIC: somatic cancer genetics at high-resolution. Nucleic Acids Res 45, D777–D783 (2017).

26 Cerami, E., Gao, J., Dogrusoz, U., Gross, B. E., Sumer, S. O., Aksoy, B. A. et al. The cBio Cancer Genomics Portal: An Open Platform for Exploring Multidimensional Cancer Genomics Data. Cancer Discov 2, 401 (2012).

27 Gao, J., Aksoy, B. A., Dogrusoz, U., Dresdner, G., Gross, B., Sumer, S. O. et al. Integrative analysis of complex cancer genomics and clinical profiles using the cBioPortal. Sci Signal 6, pl1 (2013).

28 Groschel, S., Bommer, M., Hutter, B., Budczies, J., Bonekamp, D., Heining, C. et al. Integration of genomics and histology revises diagnosis and enables effective therapy of refractory cancer of unknown primary with PDL1 amplification. Cold Spring Harb Mol Case Stud 2, a001180 (2016).

29 Kundra, R., Zhang, H., Sheridan, R., Sirintrapun, S. J., Wang, A., Ochoa, A. et al. OncoTree: A Cancer Classification System for Precision Oncology. JCO Clin Cancer Inform 5, 221–230 (2021).

30 McEvoy, C. R., Semple, T., Yellapu, B., Choong, D. Y., Xu, H., Mir Arnau, G. et al. Improved next-generation sequencing pre-capture library yields and sequencing parameters using on-bead PCR. Biotechniques 68, 48–51 (2020).

31 Chang, M. T., Bhattarai, T. S., Schram, A. M., Bielski, C. M., Donoghue, M. T. A., Jonsson, P. et al. Accelerating Discovery of Functional Mutant Alleles in Cancer. Cancer Discov 8, 174–183 (2017).

32 Alexandrov, L. B., Nik-Zainal, S., Wedge, D. C., Aparicio, S. A., Behjati, S., Biankin, A. V. et al. Signatures of mutational processes in human cancer. Nature 500, 415–421 (2013).

33 Cheng, D. T., Mitchell, T. N., Zehir, A., Shah, R. H., Benayed, R., Syed, A. et al. Memorial Sloan Kettering-Integrated Mutation Profiling of Actionable Cancer Targets (MSK-IMPACT): A Hybridization Capture-Based Next-Generation Sequencing Clinical Assay for Solid Tumor Molecular Oncology. J Mol Diagn 17, 251–264 (2015).

34 Ahmed, A. A., Etemadmoghadam, D., Temple, J., Lynch, A. G., Riad, M., Sharma, R. et al. Driver mutations in TP53 are ubiquitous in high grade serous carcinoma of the ovary. J Pathol 221, 49–56 (2010).

35 Cancer Genome Atlas, N. Genomic Classification of Cutaneous Melanoma. Cell 161, 1681–1696 (2015).

36 Cancer Genome Atlas, N. Comprehensive molecular characterization of human colon and rectal cancer. Nature 487, 330–337 (2012).

37 Li, H. & Cao, W. Pulmonary enteric adenocarcinoma: a literature review. J Thorac Dis 12, 3217–3226 (2020).

38 Network, C. G. A. R. Integrated Genomic Characterization of Pancreatic Ductal Adenocarcinoma. Cancer cell 32, 185–203.e113 (2017).

39 Arai, Y., Totoki, Y., Hosoda, F., Shirota, T., Hama, N., Nakamura, H. et al. Fibroblast growth factor receptor 2 tyrosine kinase fusions define a unique molecular subtype of cholangiocarcinoma. Hepatology 59(**4**), 1427–1434 (2014).

40 Javle, M. M., Murugesan, K., Shroff, R. T., Borad, M. J., Abdel-Wahab, R., Schrock, A. B. et al. Profiling of 3,634 cholangiocarcinomas (CCA) to identify genomic alterations (GA), tumor mutational burden (TMB), and genomic loss of heterozygosity (gLOH). J Clin Oncol 37, 4087–4087 (2019).

41 Silverman, I. M., Murugesan, K., Lihou, C. F., Féliz, L., Frampton, G. M., Newton, R. C. et al. Comprehensive genomic profiling in FIGHT-202 reveals the landscape of actionable alterations in advanced cholangiocarcinoma. J Clin Oncol 37, 4080–4080 (2019).

42 Greco, F. A., Oien, K., Erlander, M., Osborne, R., Varadhachary, G., Bridgewater, J. et al. Cancer of unknown primary: progress in the search for improved and rapid diagnosis leading toward superior patient outcomes. Ann Oncol 23, 298–304 (2012).

43 Horlings, H. M., van Laar, R. K., Kerst, J. M., Helgason, H. H., Wesseling, J., van der Hoeven, J. J. et al. Gene expression profiling to identify the histogenetic origin of metastatic adenocarcinomas of unknown primary. J Clin Oncol 26(**27**), 4435–4441 (2008).

44 Pauli, C., Bochtler, T., Mileshkin, L., Baciarello, G., Losa, F., Ross, J. S. et al. A Challenging Task: Identifying Patients with Cancer of Unknown Primary (CUP) According to ESMO Guidelines: The CUPISCO Trial Experience. Oncologist 26, e769–e779 (2021).

45 Ramaswamy, S., Tamayo, P., Rifkin, R., Mukherjee, S., Yeang, C. H., Angelo, M. et al. Multiclass cancer diagnosis using tumor gene expression signatures. Proc Natl Acad Sci U S A 98, 15149–15154 (2001).

46 Palmeri, S., Lorusso, V., Palmeri, L., Vaglica, M., Porta, C., Nortilli, R. et al. Cisplatin and gemcitabine with either vinorelbine or paclitaxel in the treatment of carcinomas of unknown primary site : results of an Italian multicenter, randomized, phase II study. Cancer 107, 2898–2905 (2006).

47 Pavlidis, N. Optimal therapeutic management of patients with distinct clinicopathological cancer of unknown primary subsets. Ann Oncol 23 **Suppl 10**, x282–285 (2012).

48 Monzon, F. A., Lyons-Weiler, M., Buturovic, L. J., Rigl, C. T., Henner, W. D., Sciulli, C. et al. Multicenter validation of a 1,550-gene expression profile for identification of tumor tissue of origin. J Clin Oncol 27, 2503–2508 (2009).

49 Herpel, E., Rieker, R. J., Dienemann, H., Muley, T., Meister, M., Hartmann, A. et al. SMARCA4 and SMARCA2 deficiency in non-small cell lung cancer: immunohistochemical survey of 316 consecutive specimens. Ann Diagn Pathol 26, 47– 51 (2017).

50 Concepcion, C. P., Ma, S., LaFave, L. M., Bhutkar, A., Liu, M., DeAngelo, L. P. et al. Smarca4 Inactivation Promotes Lineage-Specific Transformation and Early Metastatic Features in the Lung. Cancer Discov 12, 562–585 (2022).

51 Greco, F. A. & Hainsworth, J. D. Renal Cell Carcinoma Presenting as Carcinoma of Unknown Primary Site: Recognition of a Treatable Patient Subset. Clin Genitourin Cancer 16, e893–e898 (2018).

52 Overby, A., Duval, L., Ladekarl, M., Laursen, B. E. & Donskov, F. Carcinoma of Unknown Primary Site (CUP) With Metastatic Renal-Cell Carcinoma (mRCC) Histologic and Immunohistochemical Characteristics (CUP-mRCC): Results From Consecutive Patients Treated With Targeted Therapy and Review of Literature. Clin Genitourin Cancer 17, e32–e37 (2019).

53 Yakirevich, E., Perrino, C., Necchi, A., Grivas, P., Bratslavsky, G., Shapiro, O. et al. NF2 mutation-driven renal cell carcinomas (RCC): A comprehensive genomic profiling (CGP) study. J Clin Oncol 38, 726–726 (2020).

54 Wei, E. Y., Chen, Y. B. & Hsieh, J. J. Genomic characterisation of two cancers of unknown primary cases supports a kidney cancer origin. BMJ Case Rep 2015 (2015).

55 Escudier, B. Combination Therapy as First-Line Treatment in Metastatic Renal-Cell Carcinoma. N Eng J Med 380, 1176–1178 (2019).

56 Tang, T. T., Konradi, A. W., Feng, Y., Peng, X., Ma, M., Li, J. et al. Small Molecule Inhibitors of TEAD Auto-palmitoylation Selectively Inhibit Proliferation and Tumor Growth of NF2-deficient Mesothelioma. Mol Cancer Ther 20, 986–998 (2021).

57 Priestley, P., Baber, J., Lolkema, M. P., Steeghs, N., de Bruijn, E., Shale, C. et al. Pan-cancer whole-genome analyses of metastatic solid tumours. Nature 575, 210–216 (2019).

58 Yuan, Y., Shi, Y., Li, C., Kim, J., Cai, W., Han, Z. et al. DeepGene: an advanced cancer type classifier based on deep learning and somatic point mutations. BMC Bioinformatics 17, 476 (2016).

59 Jiao, W., Atwal, G., Polak, P., Karlic, R., Cuppen, E., Subtypes, P. T. et al. A deep learning system accurately classifies primary and metastatic cancers using passenger mutation patterns. Nat Commun 11, 728 (2020).

60 Abraham, J., Heimberger, A. B., Marshall, J., Heath, E., Drabick, J., Helmstetter, A. et al. Machine learning analysis using 77,044 genomic and transcriptomic profiles to accurately predict tumor type. Transl Oncol 14, 101016 (2021).

