## Supplementary Methods for "DNA sequencing and gene-expression profiling assists in making a tissue of origin diagnosis in cancer of unknown primary"

**Study Recruitment**

CUP patients were recruited to the SUPER study centralized at the Peter MacCallum Cancer Centre (PMCC) from 11 participating institutions across Australia between 2014-2020: Flinders Medical Centre, Westmead Hospital, Healthscope Pathology, Geelong Hospital - Barwon Health Andrew Love Cancer Centre, South West Healthcare Warrnambool Campus, Nepean Hospital, Blacktown Mount Druitt Hospital, Border Medical Oncology, Bendigo Health, Canberra Hospital, Royal Darwin Hospital.

Clinical data was collected at baseline, and 6- and 12-months post baseline, or until death (whichever occured first). Clinical data collected at baseline included:

- Personal details – including hospital medical record number, Medicare number (if separate consent was provided), full name, address, date of birth, contact details, next of kin or secondary contact details;
- Date of diagnosis, histopathological findings, and metastatic sites of disease;
- Diagnostic imaging reports – including chest x-ray, CT imaging, mammogram, MRI, PET, and Bone scan (if applicable);
- Pathology and any other diagnostic investigational reports;
- Primary treatment details – including surgery, radiotherapy and, chemotherapy;
- Clinician notes - including medical history, co-morbidities, previous malignancies (including treatment center), ECOG performance status, weight and weight loss; and
- Involvement with hospital services or referrals at the time of recruitment.

Clinical data collected at follow up (6 months and 12 months post baseline, or until death) includes:

- Patient status – alive and disease free, alive with disease, lost to follow up, or deceased (including date and cause of death and autopsy (if performed));
- Identification of a primary cancer, if this occurred;
- Results of any tissue of origin or molecular testing (if performed);
- Disease progression/recurrence (including date of detection) and progression free survival;
- ECOG performance status and weight;
- Resource utilization questions;
- Additional imaging, investigational or pathology reports obtained since baseline;
- Additional treatments details – including surgery, radiotherapy, chemotherapy treatments (including dates and treatment centers) since baseline;
- Changes in treatment as a result of receipt of molecular testing results (if applicable, collected at approximately the 6 month follow up time point only); and
- Involvement with hospital services or referrals since baseline.

CUPs were classified for the purpose of the study by medical oncologists into the ESMO defined clinico-pathological subsets defined as favorable and unfavorable prognosis groups (1). Briefly, favourable type includes isolated axillary tumors papillary or serous tumors in the peritoneal cavity in females, squamous carcinomas in cervical or inguinal nodes, peritoneal serous adenocarcinoma, extragonadal germ cell tumors, solitary metastases and metastatic neuroendocrine tumors. The unfavourable prognosis group present with adenocarcinomas and poorly differentiated carcinomas, often with visceral disease and multiple metastases.

Routine diagnostic formalin fixed, paraffin embedded (FFPE) tumor blocks or unstained sections were obtained from collaborating institutions. One standard EDTA tube of blood was collected from each participant for normal germline DNA. All samples were obtained according to a protocol (HREC protocol: 13/62) approved by the Peter MacCallum Cancer Centre Ethics Review Committee and consistent with Australian National Health and Medical Research Council.  Written informed consent was obtained from all participants.

Known metastatic tumors used for validation of the GEP TOO test were sourced under separate HREC approved protocol (HREC protocol 11/117). All research was conducted according to ethical standards of the Australian National Health and Medical Research Committee.

**DNA and RNA extraction**

Up to ten unstained tumor sections of five microns thickness were used for nucleic acid extraction. A hematoxylin and eosin (H&E) stained section was used to evaluate tumor cellularity, and to guide tumor macrodissection from unstained sections. The tissue was dissected and digested as previously described (2). DNA and RNA were extracted using the AllPrep DNA/RNA FFPE Kit (Qiagen, Germany) according to the manufacturer’s instructions. Germline DNA Extraction and purification of DNA was performed using the DNeasy Blood & Tissue Kit (Qiagen, Germany) spin-column procedure on the QIAsymphony (Qiagen, Germany) as per manual. DNA quantification was performed using the Qubit dsDNA HS Assay kit and the Qubit 2.0 Fluorometer (Life Technologies, Carlsbad, CA, USA). RNA recovery was measured using NanoDrop 3300 Fluorometer (Thermo Scientific, Waltham, MA, USA).

**Gene expression** **profiling**

We used two different GEP tests for TOO classification. A previously described microarray-based classifier (CUPGuide) (3) which was discontinued as a commercial test during the recruitment phase, leading to the development of a second classifier using the nCounter molecular barcode multiplexed assay (NanoString Technologies Inc.), performed according to the manufacturer's guidelines. NanoString involved 225 genes differentially expressed across 18 tumor classes identified from the original training dataset for CUPGuide. Like CUPGuide, each class in the NanoString model represented a broad cancer class based on TOO with the exception of SCCs, neuroendocrine neoplasms, and sarcomas that were pooled together into their respective classes. In addition to genes selected for TOO classification, the NanoString panel included probes for six endogenous control genes and three oncoviral transcripts (HPV16, HPV18, MCPyV). Other cancer and immune-related genes were included on the panel but not used for classification (Supplementary Table x).

Detection of viral transcripts for HPV16 L1 major capsid protein, HPV18 L1 major capsid protein and Merkel cell polyomavirus capsid protein VP2 reported where raw transcript counts are detected above background levels relative to the NanoString reference set (>50 transcript counts).

**Gene expression normalization**

A cross-platform TOO classifier was trained on RNA-sequencing data representing 8,454 primary and metastatic tumors from 18 cancer types from the TCGA. A k-nearest neighbors (kNN) with Pearson correlation distance metric on TCGA reference set was used to classify an independent validation set consisting of 188 known origin metastatic tumors profiled on the NanoString panel. The final classifier utilized 10 neighbors that produces overall accuracy of 82.98%, with consideration of per-class accuracy (refer to Supplementary Table xx. full breakdown of accuracy/specificity), as well as principle of parsimony. A probability score was generated for kNN testing and heuristic thresholds set for classification confidence level (unclassified <0.5, low ≥ 0.5 and ≤ 0.7, medium confidence >0.7 and < 0.9, high confidence ≥9 probability). The 188 known origin metastatic tumors were used as a reference set for CUP samples. The NanoString gene expression raw count data was normalized using the geometric mean of housekeeping genes from the reference set. The CUP and reference set normalized data was then further transformed using  Z-score standardization.

R (version 3+) scripts were used to build and train the classifier and available in the public project GitHub repository.

**Targeted DNA sequencing**

Targeted sequence analysis was performed on matched blood and tumor DNA. Two hybridization targeted capture panels were used  to screen for mutations in the coding regions and splice sites of 626 cancer-related genes in the first iteration of the assay and 386 cancer-related genes in the second and most recent iteration of the assay (list of genes Supp Table 3). DNA libraries were prepared using a KAPA Library Preparation Kit (Roche Holdings, Inc., South San Francisco, CA, USA) and enriched using a SureSelect XT Target Enrichment assay (Agilent Technologies, Santa Clara, California, CA, USA) (Agilent Design ID 0628271 for version 1 and Design ID 3016871 for version 2). Full methods of the panel has been previously described (4).

**DNA sequencing variant calling and analysis**

Alignment and variant calling was performed using the ensemble variant caller bcbio-nextgen (BCBio) cancer somatic variant calling pipeline (version 1.1.3a) ([https://github.com/bcbio/bcbio-nextgen](https://github.com/bcbio/bcbio-nextgen%22%20/t%20%22_blank)) and aligned using human genome 38 reference. BAM and Fastq files from both tumor and matched germline were analyzed through the BCBio pipeline for detection of small nucleotide variant (SNV) mutations. To maintain consistency between the two iterations of the targeted panel, variant calling from the DNA sequencing results were restricted to the coverage and panel size of the second version of the panel (2.024 Mb).

SNVs and Indels were interpreted using Personal Cancer Genome Reporter (5) and filtered based on the variant Tier level of clinical significance using the adapted structure of Association for Molecular Pathology, American Society of Clinical Oncology, and College of American Pathologists (6). Mutations that were annotated as clinical significance (Tier 1), potential clinical significance (Tier 2) or variants of uncertain significance (Tier 3) were included, and non-coding variants (Tier 4) were removed. Variants were further selected based on their tumor purity and variant allele frequency (VAF). A VAF threshold of above 5% was used in tumors with sufficient tumor purity fraction (>=20%). In tumors with a low tumor purity (<20%) but with mutations that were considered reliable, verified with supporting reads and clinically important, a VAF of 1% was used as a cutoff.

The R tool *facets* was used to estimate tumor purity and ploidy as well as detected copy number alterations in the tumor sample ([https://github.com/mskcc/facets](https://github.com/mskcc/facets%22%20/t%20%22_blank)) (7). Genes were considered amplified if the copy number was three times greater than the ploidy value or homozygous deletion if the copy number was zero. Structural variants (SVs) were detected using the R package *gridss* (8). SV genes were limited to those of potential clinical importance, specifically, *FGFR2, FGFR3, ERG, TMPRSS2, ALK, RET, ROS1, NRG1, NTRK1*. The R package *maftools* ([https://github.com/PoisonAlien/maftools](https://github.com/PoisonAlien/maftools%22%20/t%20%22_blank)) (9) was used to visualize the variants called by BCBio with the filtering criteria described above.

Cosmic version 2 mutational signatures were called using the R package *MutationalPatterns* ([https://github.com/UMCUGenetics/MutationalPatterns](https://github.com/UMCUGenetics/MutationalPatterns%22%20/t%20%22_blank)) with the reference Bioconductor genome BSgenome.Hsapiens.UCSC.hg38. Due to the limited size of the targeted panel, a contribution of over 50 mutations were required to call a mutational signature and the signature had to contribute to at least 25% of mutational processes. Signatures investigated were reduced to those of clinical utility and typically of high mutation load, specifically, COSMIC V2 signatures 4, 6, and 7.

DNA capture bait libraries did not target HPV and EBV but viral sequences were detected in off-target sequencing reads of the tumor DNA BAM files using pathogen detection tools  *Xenomapper* ([https://github.com/genomematt/xenomapper](https://github.com/genomematt/xenomapper%22%20/t%20%22_blank)) and *oviraptor* ([https://github.com/vladsaveliev/oviraptor](https://github.com/vladsaveliev/oviraptor%22%20/t%20%22_blank)). Oncovirus sequences for HPV16, HPV18, MCPV and EBV were investigated, the former three were validated with NanoString virus transcripts (accession IDs NC_001526, U89349, NC_010277.2 and NC_007605), while EBV detection was based on DNA sequencing alone (accession NC_007605) as EBV probes were not included in the NanoString assay.

Tumor mutation burden was calculated by dividing the total number of coding mutations detected by BCBio by the coverage of the respective panels (V1 2.999 Mb and V2 2.024Mb).

References

1.	Fizazi K, Greco FA, Pavlidis N, Daugaard G, Oien K, Pentheroudakis G, et al. Cancers of unknown primary site: ESMO Clinical Practice Guidelines for diagnosis, treatment and follow-up. Ann Oncol. 2015;26 Suppl 5:v133-8.

2.	Wong SQ, Fellowes A, Doig K, Ellul J, Bosma TJ, Irwin D, et al. Assessing the clinical value of targeted massively parallel sequencing in a longitudinal, prospective population-based study of cancer patients. Br J Cancer. 2015;112(8):1411-20.

3.	Tothill RW, Li J, Mileshkin L, Doig K, Siganakis T, Cowin P, et al. Massively-parallel sequencing assists the diagnosis and guided treatment of cancers of unknown primary. J Pathol. 2013;231(4):413-23.

4.	McEvoy CR, Semple T, Yellapu B, Choong DY, Xu H, Mir Arnau G, et al. Improved next-generation sequencing pre-capture library yields and sequencing parameters using on-bead PCR. Biotechniques. 2020(1940-9818 (Electronic)).

5.	Nakken S, Fournous G, Vodák D, Aasheim LB, Myklebost O, Hovig E. Personal Cancer Genome Reporter: variant interpretation report for precision oncology. (1367-4811 (Electronic)).

6.	Li MM, Datto M, Duncavage EJ, Kulkarni S, Lindeman NI, Roy S, et al. Standards and Guidelines for the Interpretation and Reporting of Sequence Variants in Cancer: A Joint Consensus Recommendation of the Association for Molecular Pathology, American Society of Clinical Oncology, and College of American Pathologists. 2017(1943-7811 (Electronic)).

7.	Shen R, Seshan VE. FACETS: allele-specific copy number and clonal heterogeneity analysis tool for high-throughput DNA sequencing. Nucleic Acids Res. 2016;44(16):e131.

8.	Cameron DL, Schröder J, Penington JS, Do H, Molania R, Dobrovic A, et al. GRIDSS: sensitive and specific genomic rearrangement detection using positional de Bruijn graph assembly. Genome research. 2017;27(12):2050-60.

9.	Mayakonda A, Lin DC, Assenov Y, Plass C, Koeffler HP. Maftools: efficient and comprehensive analysis of somatic variants in cancer. Genome Res. 2018;28(11):1747-56.
