## Supplementary figures and images for "DNA sequencing and gene-expression profiling assists in making a tissue of origin diagnosis in cancer of unknown primary"

### Supplementary Figure 1

#### Supplementary Figure 1

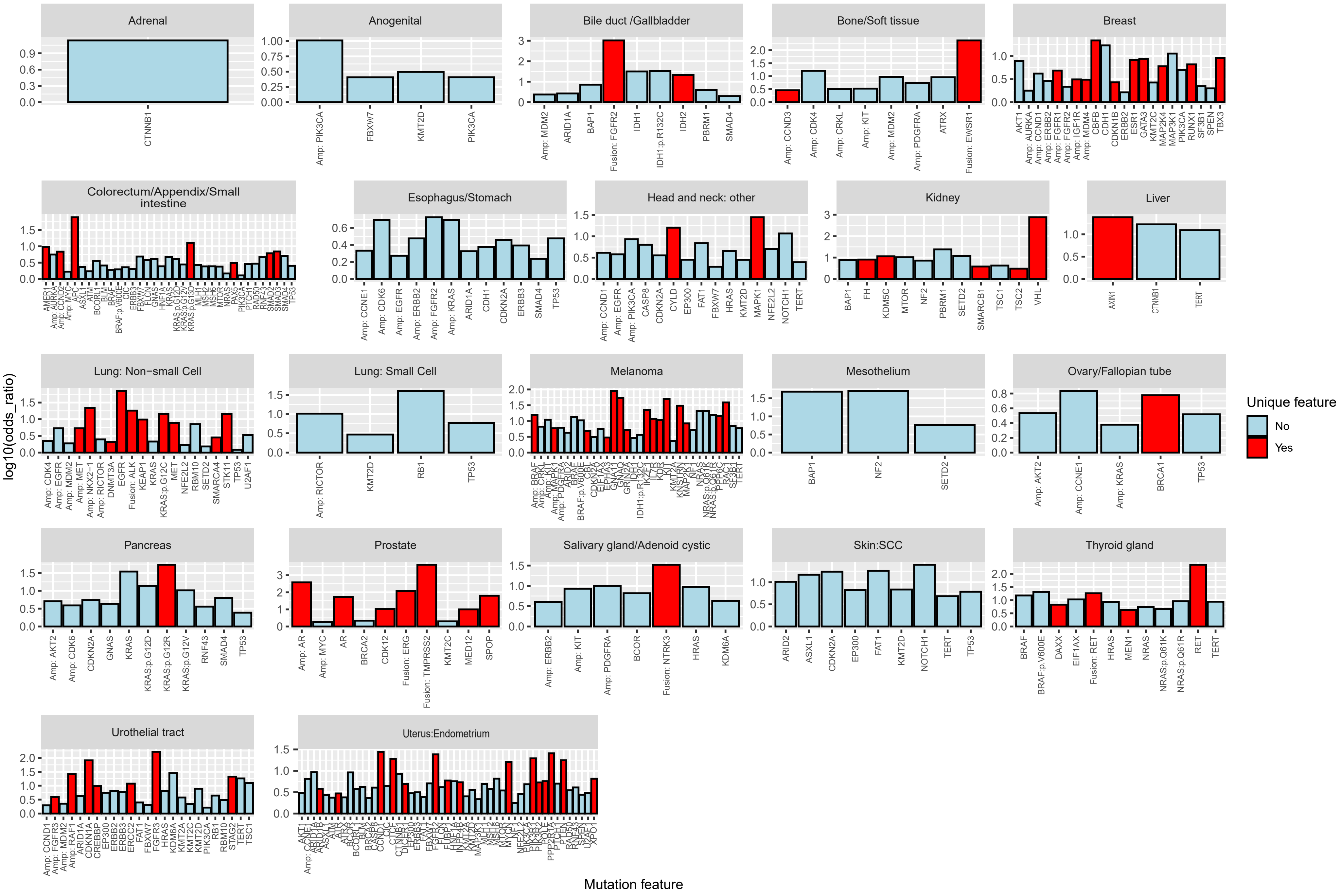

### Supplementary Figure 2

#### Supplementary Figure 2

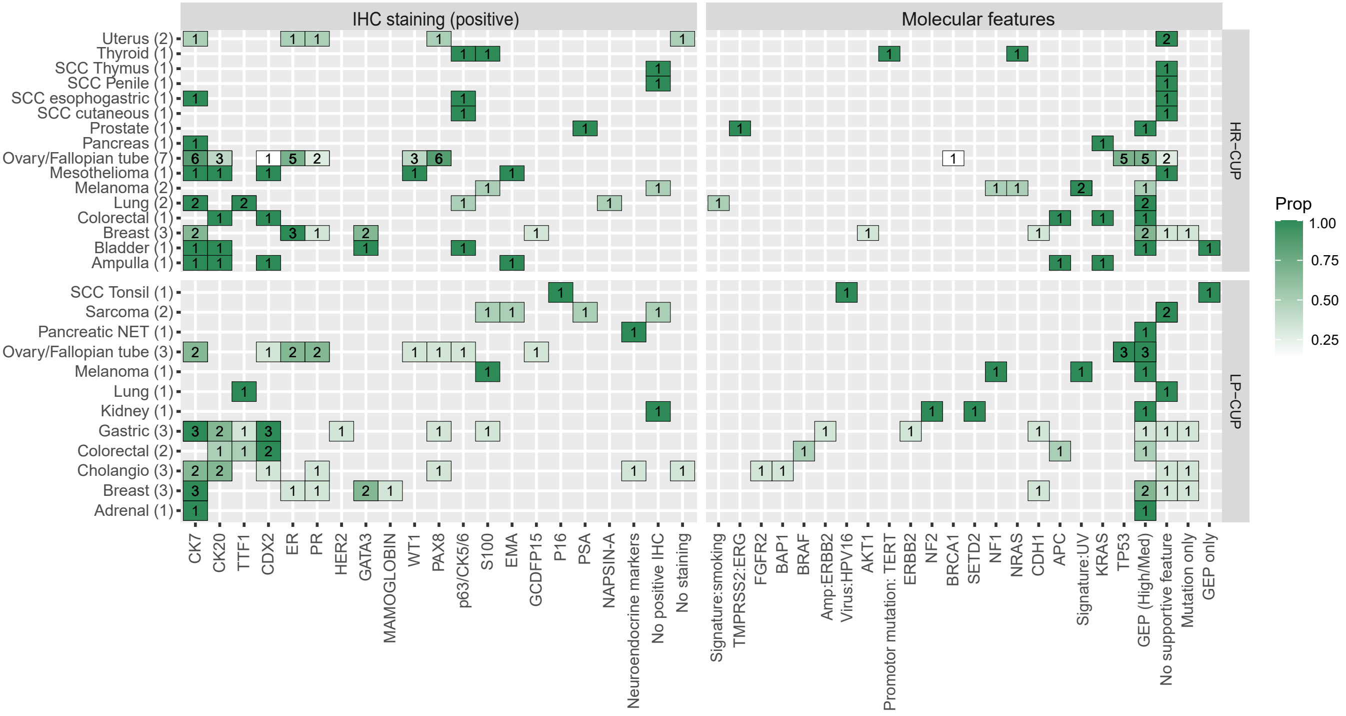
